# Detecting the Emergent or Re-Emergent COVID-19 Pandemic in a Country: Modelling Study of Combined Primary Care and Hospital Surveillance

**DOI:** 10.1101/2020.05.13.20100743

**Authors:** Nick Wilson, Markus Schwehm, Ayesha J Verrall, Matthew Parry, Michael G Baker, Martin Eichner

## Abstract

**Aims:** We aimed to determine the effectiveness of surveillance using testing for SARS-CoV-2 to identify an outbreak arising from a single case of border control failure at a country level.

**Methods:** A stochastic version of the SEIR model CovidSIM v1.1 designed specifically for COVID-19 was utilised. It was seeded with New Zealand (NZ) population data and relevant parameters sourced from the NZ and international literature.

**Results:** For what we regard as the most plausible scenario with an effective reproduction number of 2.0, the results suggest that 95% of outbreaks from a single imported case would be detected in the period up to day 33 after introduction. At the time point of detection, there would be a median number of 6 infected cases in the community (95%UI: 1–68). To achieve this level of detection, an on-going programme of 7,800 tests per million people per week for the NZ population would be required. The vast majority of this testing (96%) would be of symptomatic cases in primary care settings and the rest in hospitals. Despite the large number of tests required, there are plausible strategies to enhance testing yield and cost-effectiveness eg, (i) adjusting the eligibility criteria via symptom profiles; (ii) and pooling of test samples.

**Conclusions:** This model-based analysis suggests that a surveillance system with a very high level of routine testing is probably required to detect an emerging or re-emerging SARS-CoV-2 outbreak within one month of a border control failure in a nation.

## Introduction

One of the challenges with a new pandemic such as COVID-19, is how best to undertake surveillance. Good quality surveillance is needed to maximise rapid disease control eg, with case isolation and contact tracing to identify further cases and to quarantine contacts. This capacity is particularly critical for nations that decide to contain spread at an early stage and even eliminate transmission entirely (eg, as New Zealand has planned [1] and Australia has as a potential option [2]). In particular it may be relevant to the following groupings of island jurisdictions (as per WHO data on 5 May 2020 [3]):

- Those jurisdictions which have avoided any COVID-19 cases to date (eg, via border controls), but which are still at risk if border controls fail. These mainly include island jurisdictions in the Pacific Ocean (eg, American Samoa, Cook Islands, Federated States of Micronesia, Kiribati, Marshall Islands, Nauru, Niue, Palau, Samoa, Solomon Islands, Tokelau, Tonga, Tuvalu, Vanuatu, and Wallis and Futuna).
- Those jurisdictions which have only experienced sporadic cases and may have successfully contained spread. These are mainly Caribbean islands (eg, Trinidad and Tobago) but also include some islands in the Pacific (eg, Fiji).
- Those jurisdictions which have had larger outbreaks of COVID-19, but have instituted tight border controls and have declining numbers of new cases. These settings may therefore be on track towards eliminating the virus (eg, Australia, New Zealand and Taiwan).

A recent Australian study [4], suggested that timely detection and management of community transmission of COVID-19 is feasible. This modelling study concluded that “testing for infection in primary care patients presenting with cough and fever is an efficient, effective and feasible strategy for the detection and elimination of transmission chains”. For example, when testing 9000 people per week (per million population), the authors estimated that no cases of COVID-19 would be missed in some circumstances. This type of surveillance could therefore be relevant to identifying emergent or re-emergent SARS-CoV-2, the pandemic virus causing COVID-19.

Given this background, we aimed to determine the effectiveness of surveillance using testing for the SARS-CoV-2 virus to identify an outbreak arising from a single case of border control failure in a nation without it.

## Methods

To run pandemic spread scenarios for New Zealand, we used a stochastic SEIR type model with key compartments for: susceptible [S], exposed [E], infected [I], and recovered/removed [R]. The model is a stochastic version of CovidSIM which was developed specifically for COVID-19 by two of the authors (http://covidsim.eu; version 1.1). Work has been published from version 1.0 of the deterministic version of the model [5] [6], but in the Appendix we provide updated parameters and differential equations for version 1.1. The stochastic model was built in Pascal and 100,000 simulations were run for each set of parameter values.

The parameters were based on available publications and best estimates used in the published modelling work on COVID-19 (as known to us on 6 May 2020). Key components were: a single undetected infected case arriving in New Zealand via a border control failure, 80% of infected COVID-19 cases being symptomatic, 39.5% of cases seeking a medical consultation in primary care settings, and 4% of symptomatic cases being hospitalised (see Table A1 in the Appendix for further details). We assumed that the initial undetected case could be at any stage of infection – to cover both failures of managing quarantine at the border, but also failures around the management of non-quarantined workers such as air-crew and ship-crew. Scenarios considered different levels of transmission with the effective reproduction number (Re) of SARS-CoV-2 to be 1.5, 2.0, 2.5 and 3.0 (Table A1). Other scenarios considered the impact of 75% of symptomatic people seeking a medical consultation (eg, as the result of a potential media campaign); and another considered a possible school outbreak (eg, a border control failure involving a teacher or student returning from overseas). The assumptions for the latter involved: Re = 2.0, only 5% of symptomatic cases seek medical consultation, and only 0.5% being hospitalised.

For the detection of COVID-19 cases, we assume testing of 95% of cases of symptomatic cases of respiratory illness seeking medical attention in primary care and of hospitalised cases of respiratory illness. For parameterising the size of these two groups, we used official statistics and results from the Flutracking surveillance system used in New Zealand (Table A1). The sensitivity of the PCR diagnostic test (at 89%) was based on a meta-analysis (Table A1).

## Results

For what we regard as the most plausible scenario with an Re of 2.0 (ie, where people are practicing some level of reduced social contact because of the pandemic in other countries), the results suggest that 50% of outbreaks from a single imported case would be detected in the period up to day 13 and 95% in the period up to day 33 (Table 1, Figure 1). At the time of detection (to day 33), there was an estimated median number of 6 infected cases in the community (95% uncertainty interval [UI]: 1− 68). Similarly, for this same period, we expected that 4.7 primary care consultations and 0.5 hospitalisations of COVID-19 patients will have occurred. To achieve this level of detection, an on-going programme of 5,600 tests per day would be required, which is 7,800 tests per million people per week for the whole New Zealand population. This is one test per 128 individuals per week when combining the testing of the 39.5% seeking medical attention for cough and fever symptoms and the testing of those being hospitalised for respiratory conditions. The vast majority of this testing (96%) would however, be in primary care settings and the rest in hospitals.

**Table 1:** Modelled impacts by the time it takes to obtain at least one positive test result for SARS-CoV-2 arising from a border control failure where a single case enters the island nation of New Zealand (all results adjusted for lag times in reporting and obtaining test results, using 100,000 stochastic simulations for each parameter setting) [Nick move tests to end].

| Scenario with variation in the effective reproduction number (Re) | Introduction leads to no further spread (%) | Day when 50% of outbreaks have been detected (median) | Day when 95% of outbreaks have been detected (median) | Mean day of outbreak detection | Expected no. of total tests* | Median (95% UI) number of infections from introduction to detection** | Expected cumulative primary care consultations for COVID-19 from introduction to detection | Expected cumulative hospitalisations for COVID-19 from introduction to detection |
| --- | --- | --- | --- | --- | --- | --- | --- | --- |
| Re = 1.5 | 42.8 | 13 | 35 | 15.2 | 84,800 | 4 (1-36) | 2.9 | 0.3 |
| Re = 2.0 (most plausible) | 35.3 | 13 | 33 | 15.0 | 83,700 | 6 (1-68) | 4.7 | 0.5 |
| Re = 2.5 | 30.1 | 13 | 30 | 14.5 | 80,900 | 9 (1-111) | 7.4 | 0.8 |
| Re = 3.0 | 26.2 | 13 | 28 | 14.1 | 78,700 | 12 (1-167) | 10.1 | 1.0 |
| Re = 1.5 but 75% seek medical consultation | 35.5 | 9 | 24 | 10.4 | 106,000 | 3 (1-20) | 3.1 | 0.2 |
| Re = 2.0 but 75% seek medical consultation | 30.6 | 10 | 23 | 10.6 | 108,000 | 3 (1-36) | 4.8 | 0.3 |
| School outbreak*** | 41.5 | 32 | 65 | 33.0 | 184,000 | 47 (2-562) | 4.9 | 0.5 |

\* From the time of the border control failure to the mean day of outbreak detection. This is around 5,600 tests per day for primary care and hospital sectors combined for the first four listed scenarios.
\*\* Includes those in the latent phase, prodromal phase, asymptomatic infections and symptomatic infections (but not recovered or deceased cases).
\*\*\* The assumed characteristics for this school outbreak involved: Re = 2.0, only 5% of symptomatic cases seeking medical consultation, and only 0.5% being hospitalised. The level of testing was as per the first four listed scenarios.

For all scenarios, 95% of outbreaks were detected before five weeks after introduction. The highest value (32 days) was for the simulated school outbreak where medical consultations were assumed to be much less likely (due to symptoms in young people being typically milder). Increasing the extent by which symptomatic people seek medical consultations to the 75% level (up from that reported by Flutracking at 39.5%), would reduce the time to detection (eg, from 33 to 23 days for the Re = 2.0 scenario at the 95% probability level, Table 1).

**Figure 1:**
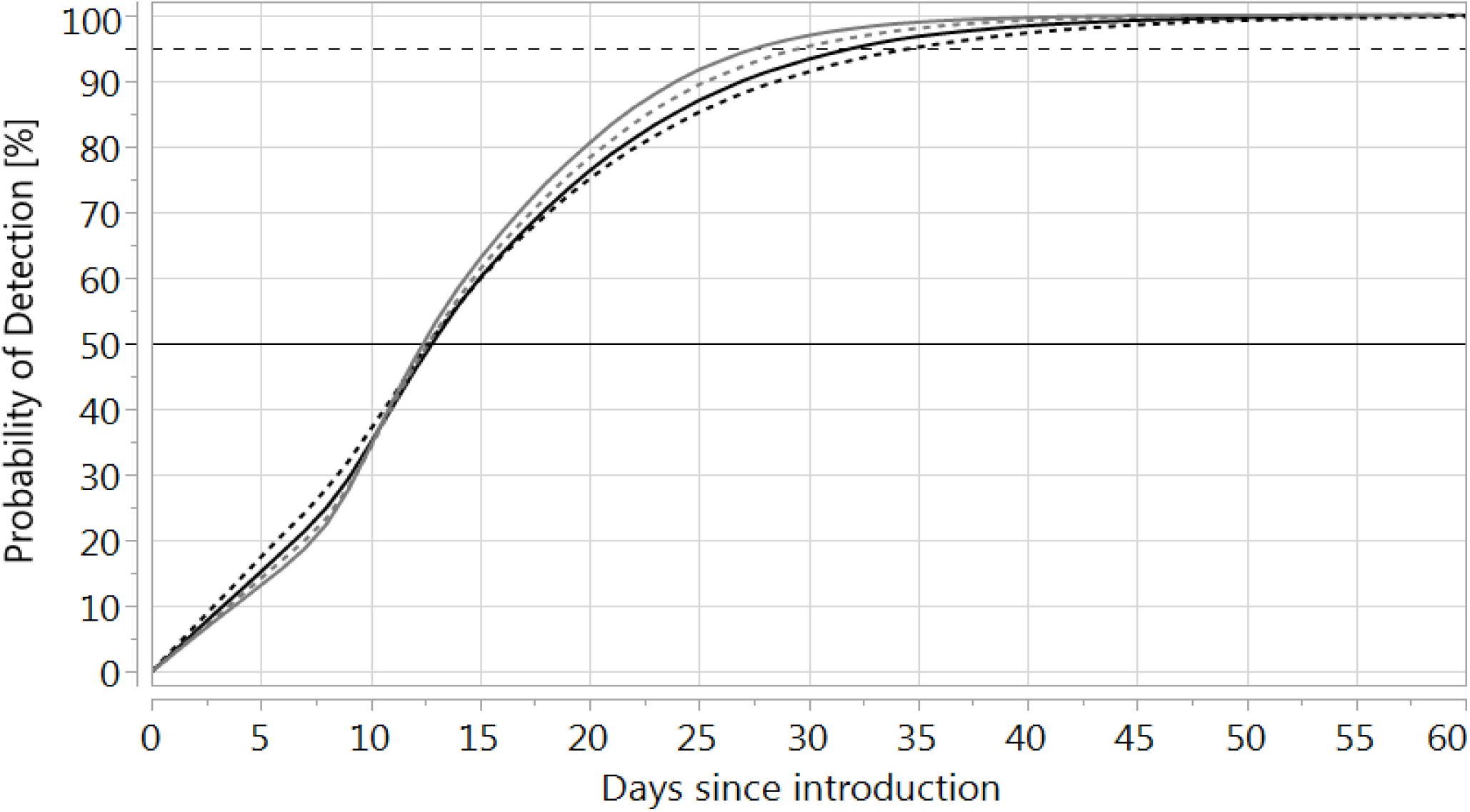
Probability of COVID-19 case detection after reintroduction of the infection (the different curves represent the results of 100,000 simulations each, using Re = 1.5 (black dotted), 2.0 (black solid), 2.5 (grey dotted), and 3 (grey solid), respectively.

## Discussion

This analysis indicates the challenges for a surveillance system designed to detect the emergence or re-emergence of SARS-CoV-2 transmission in a nation with border controls. A very high level of testing is typically required in primary care settings and hospitals to detect an outbreak arising from a single border control failure.

This testing level, at 7,800 per week per million population, is somewhat higher than the levels in NZ in early May 2020 (ie, the 7-day rolling average at this time was around 4,200 tests per day [7], which is around 6000 people per week per million population, although this included some screening of asymptomatic people). Also achieving this high level of testing would require maintaining public support and participation with testing in an environment where border control failures might be considered to be rare events (though even a 14-day quarantine period for arriving travellers is likely to have a small failure rate [8]).

Despite the high level of testing required for this type of surveillance system, there are potential ways that might improve the yield and cost-effectiveness of such testing:

- Further refining eligibility criteria for testing in the primary care setting (eg, rather than just cough and fever). This could improve both the sensitivity and specificity of the surveillance system (eg, patients with self-reported olfactory and taste disorders had high specificity as a screening criterion in a Singapore study [9]). While PCR tests typically have very high specificity, the test kits with the best characteristics could be evaluated for cost-effectiveness.
- Pooling samples for PCR testing may preserve reagents and be more cost-effective [10], but would need to be balanced against potential loss of sensitivity and associated diagnostic delays.

Furthermore, extremely comprehensive testing of *all* hospitalised cases with respiratory symptoms has some other advantages. It assists with appropriate diagnosis and management of COVID-19 cases when these are identified, but it also may protect against a hospital outbreak of COVID-19 which could endanger lives of inpatients and put many health workers into quarantine.

## Study strengths and limitations

This is the first such modelling analysis for a country with an elimination goal for COVID-19. Nevertheless, this work could have been refined further by a focus on a narrower range of acute respiratory diseases (eg, excluding the category of hospital admissions for chronic lower respiratory diseases (ICD10 codes: J40-J47). But since hospital admissions for these often involve an acute aspect eg, acute bronchitis on top of chronic obstructive respiratory disease, we took the parsimonious approach of considering all respiratory diseases.

This analysis also did not explore other surveillance options such as routine active surveillance of specific groups (eg, air-crew, ship-crew and port workers), or the testing of town and city sewerage systems for the virus, as is being explored in several jurisdictions internationally [11] [12]. Yet these surveillance issues should also be considered by nations that aim to keep the COVID-19 pandemic out or eliminate it.

## Conclusions

In conclusion, this model-based analysis suggests that a surveillance system with a very high level of routine testing is probably required to detect an emerging or re-emerging SARS-CoV-2 outbreak within one month of a border control failure in a nation. But further work is required to improve on this type of analysis and to evaluate other potential surveillance system components.

## Data Availability

Simulation results are available on request to the authors

## Appendix: Mathematical description of the CovidSIM model (version 1.1) and model parameters

The stochastic simulations are based on the following differential equations:

### Model dynamics

Number of susceptible individuals 
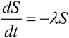

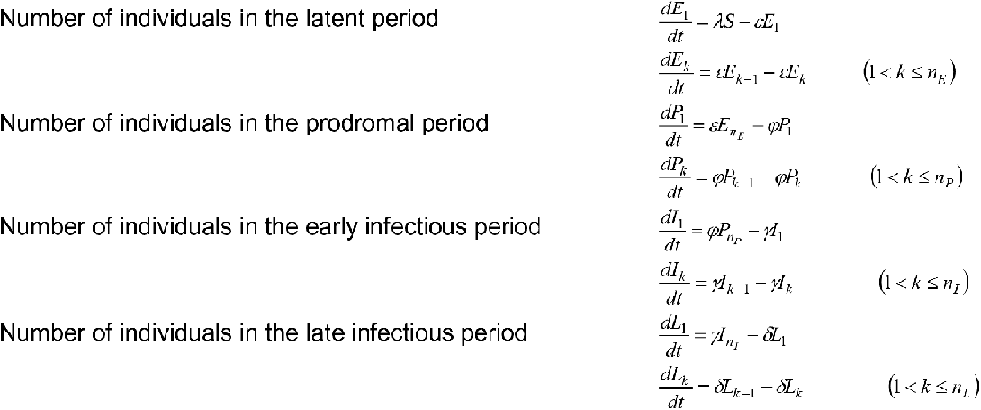

### Derived variables

Total number in latent period 
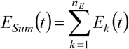

Total number in prodromal period 
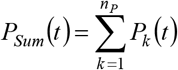

Total number in early infectious period 
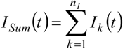

Total number in late infectious period 
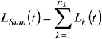

### Contact rate and force of infection

Contact rate 
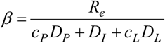

Force of infection 
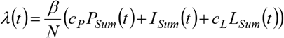

### Stochastic treatment of the differential equations

The kind of epidemiologic events and the duration between two consecutive events are calculated using random numbers. The simulations start with a fully susceptible population in which one individual (index case) is infected. The infection stage of the index case is picked at random, taking into consideration the different lengths of the latent, prodromal, early and late infectious period. In each simulation, the sum of all the rates that change the current state of the system is calculated as

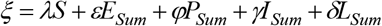

A uniformly distributed random number *r*_1_∈[0,*ξ*] is then chosen, and the time Δ*t* = −ln(*r*_1_)/*ξ* after which the next event occurs is calculated. All transition rates are arranged in an arbitrary order, and cumulative rates are calculated by adding their individual rates. A new uniformly distributed random number *r*_2_ ∈[0,*ξ*] is chosen, and the first transition in the order whose cumulative rate is larger than *r*_2_ is performed. If, for example, the event is an infection, one individual is removed from the group of susceptible individuals and added to the group of latent individuals of stage 1. New rates are calculated after each step, and the procedure is repeated. A more detailed description of the transformation of differential equation models to stochastic models can be found in Gillespie (1976) [13].

### Parameters

*N* Population size

*λ* Force of infection

*R_e_* Effective reproduction number

*β* Effective contact rate

*D_E_* Average duration of the latent period

*n_E_* Number of stages for the latent period

*ε* Stage transition rate for the latent period(*ε* = *n*_E_ / *D_E_*)

*D_P_* Average duration of the prodromal period

*n_P_* Number of stages for the prodromal period

*φ* Stage transition rate for the prodromal period (*φ=n_p_/D_p_*)

*c_P_* Contagiousness in the prodromal period (relative to the contagiousness in the early infectious period)

*D_I_* Average duration of the early infectious period

*n_I_* Number of stages for the early infectious period

*γ* Stage transition rate for the early infectious period (γ= *n_I_*/*D_I_*)

*D_L_* Average duration of the late infectious period

*n_L_* Number of stages for the late infectious period

*δ* Stage transition rate for the late infectious period (*δ* = *n_L_*/*D_L_*)

*c_L_* Contagiousness in the late infectious period (relative to the contagiousness in the early infectious period)

---

**Table A1:** Input parameters used for modelling the potential spread of the COVID-19 pandemic with the stochastic version of CovidSIM (v1.1) with New Zealand as a case study.

| Parameter | Value/s used | Further details for parameter inputs into the modelling |
| --- | --- | --- |
| Population size | 4,951,500 | We used the estimated NZ population as per December 2019 (ie, 4,951,500 [14]). |
| Infections that lead to sickness | 80% | We used the same proportion (80%) of symptomatic cases as per a Chinese study [15], and as per an Australian modelling study.[4] This value is higher than the 57% value found in an Icelandic study [16] but this study did not involve long-term follow-up of the asymptomatic cases (ie, some of the asymptomatic cases might subsequently have developed symptoms). But it is also lower than that found in another Chinese study (at 94% symptomatic) [17]. |
| Sick people who seek medical attention in primary care | 39.5% (75% in scenario analyses) | We used the result from the NZ Flutracking surveillance system for people with “fever and cough” in the weekly surveys who report seeking medical attention for these symptoms [18]. This is very similar to international estimates for people with influenza who seeking medical attention at 40% eg, as used in other modelling [5]. In scenario analyses we raised this to 75% on the assumption that a media campaign could encourage attendance for relatively mild respiratory symptoms. |
| Sick people need hospitalisation | 4% | At the time of writing on 3 May 2020, there were 8 people hospitalised in NZ with COVID-19 (out of a total of 201 actively infected cases, ie, 4.0% [19]). Of note is that modellers in the United Kingdom (UK) have used 4.4% (of all infected cases) [20], and for modelling in the United States 3%, 5% and 12% have been proposed [21]. The length of hospitalisation was assumed to be 10 days which is similar to other modelling work eg, 10.4 days for the UK [20]. |
| Effective reproduction number ($R_e$) | 2.0 as the most plausible value for NZ (1.5, 2.5, and 3.0 used in scenario analyses) | This estimate of 2.0 is in the lower end of the range for the basic reproduction number ($R_0$) reported on 6 March by the WHO (ie, 2.0 to 2.5 [22]). This is because we assumed some level of ongoing physical distancing and enhanced hygiene practices in NZ relative to the pre-pandemic world. Of note is that an earlier review of 12 studies [23], suggested estimates that ranged from 1.4 to 6.49, with a mean of 3.28, a median of 2.79 and interquartile range of 1.16. UK modelling work has used an estimate of 2.4 (range: 2.0 to 2.6) [20]. Australian modelling studies have used $R_0$ values in the 2.2 to 2.7 range [2]. |
| Relative contagiousness in the prodromal period | 50% | There is uncertainty around this value but we used the same estimate as in recent UK modelling [20]. This has biological plausibility as while there is similarity in viral loads between asymptomatic and symptomatic COVID-19 patients [24], it would be expected that those who are fully symptomatic (with a cough etc.) would be more likely to transmit infection. Of note is an estimate from the Diamond Princess cruise ship outbreak, that 17.9% of COVID-19 infections were from asymptomatic individuals (95% credible interval 15.5-20.2%) [25]. But it is unclear how generalisable this finding is given the crowded cruise ship conditions and the typically elderly nature of the passengers. |
| Latency period | 4 days | We used an average duration of 4 days as per Read et al [26], with a standard deviation (SD) of 25% (1 day) (calculated using 16 stages; Erlang distribution). This is similar to the estimate in a Chinese study which reported a median latent period of 3.69 days [27]. |
| Prodromal period | 1 day | There is as yet insufficient data on this for COVID-19, so we used an assumed value for influenza (SD = 25%; 0.25 days), Erlang distribution). |
| Symptomatic period | 10 days (split into 2 periods of 5 days each) | The WHO-China Joint Mission report stated that “the median time from onset to clinical recovery for mild cases is approximately 2 weeks and is 3-6 weeks for patients with severe or critical disease” [28]. But given that mild cases may have been missed in this particular assessment, we used a |
|  |  | slightly shorter total time period of 10 days (SD = 25%; 2.5 days), Erlang distribution). |
| Contagiousness during the two symptomatic periods | 100% and 50% | In the first five days of symptoms, cases were considered to be fully contagious. In the second five-day period, this was assumed to be at 50%. The latter figure is highly uncertain, but is broadly consistent with one study on changing viral load [29]. |
| **Provision of testing and test sensitivity assumed** |  |  |
| Background hospital admissions for respiratory conditions in NZ | 234 admissions per day | Using 85,439 respiratory disease admissions to NZ public hospitals in the year 2016/2017 (for all Chapter X ICD10 codes: J00 to J99) [30]. |
| Background medical consultations in primary care for respiratory conditions in NZ | 5,640 consultations per day | Data from the NZ arm of the Flutracking surveillance system was used. This indicates that approximately 3% of respondents in the period from April to October report “fever and cough” in the weekly surveys [18]. Of these 39.5% report seeking medical attention for their symptoms. However, we assumed a lower annual rate of 2% to account for the period outside of the influenza season (eg, Flutracking reporting is closer to 1% for weekly “fever and cough” at the start of May when the surveillance system begins for the year). In the NZ population of 5 million this would suggest 14,300 new cases developing “cough and fever” per day of whom 5640 would be expected to seek medical attention. |
| Coverage in patients with respiratory symptoms who seek medical attention in primary care | 95% coverage | These coverage values were further adjusted for the test sensitivity of 89% (see below). With 95% coverage and 89% test sensitivity, 84.55% of cases would be detected. |
| Coverage in hospitalised patients with respiratory symptoms | 95% coverage | As above. |
| PCR test sensitivity | 89% | A meta-analysis has reported this as 89% (95%CI: 81 to 94%) [31]. This sensitivity is not ideal as while infection can be in the lungs, the sampling is from the nasopharynx, which may contain lower levels of virus at some stages of infection. Specificity is close to 100% for the PCR test. |
| **Lag times (for health sector interaction and testing delays)** |  |  |
| Delay from symptom onset until a test has been performed and the result has become available | 5 days plus 1 day for the testing delay | There is a delay between symptom onset and the performance of the test for SARS-CoV-2. For the first part of the delay we considered a study in Beijing, China, which reported the interval time from between illness onset and seeing a doctor was 4.5 days [32]. Another Chinese study of 710 patients with pneumonia [33] reported that those dying had a median duration from onset of symptoms to radiological confirmation of pneumonia of 5 (IQR: 3–7) days.<br>For the testing delay we noted that the aim in NZ is to obtain the result of the tests in under 24-hours regardless of the primary care or hospital setting. But this may not always be the case for rural and small town settings. In our simulations, test results were available on average 5.94 days after symptom onset (SD: 1.36 days). |

